# Association of a polygenic risk score with coronary atherosclerotic burden in clinical CT angiograms

**DOI:** 10.64898/2026.05.26.26353801

**Authors:** Katherine Hartmann, Michael Gannon, Pradeep Natarajan, Philip Greenland, Penn Medicine Biobank, Michael G. Levin

## Abstract

**Background:** Polygenic risk scores (PRS) for coronary artery disease (CAD) are associated with cardiovascular events, but the relationship between inherited risk and routinely reported coronary computed tomography angiography (CTA) findings has not been studied.

**Objectives:** To evaluate associations between a genome-wide PRS for angiographic coronary disease burden and coronary CTA-derived measures of atherosclerotic severity in a real-world clinical cohort.

**Methods:** We studied Penn Medicine BioBank participants with available genotypes and clinically obtained coronary CTA reports. A previously published PRS for angiographic CAD burden was calculated using pgsc_calc. CAD-RADS scores and coronary artery calcium (CAC) values were extracted from radiology reports using the large language model Llama 3.1 8B. Associations between PRS and CAD-RADS severity were evaluated using Bayesian cumulative ordinal logit regression, while associations with log-transformed CAC burden were assessed using Bayesian linear regression.

**Results:** Among 630 participants, median age was 59 years (IQR 49 - 68), 53% were female, 62% were genetically similar to a European reference population, and 34% to an African reference population. LLM-extracted CAD-RADS and CAC values demonstrated near-perfect agreement with manual abstraction. Higher PRS was associated with greater coronary atherosclerotic burden on CTA. Each 1-standard deviation (SD) increase in PRS was associated with a 20% higher odds of belonging to a more severe CAD-RADS category (cumulative OR 1.20, 95% credible interval 1.06-1.44). Higher PRS was also associated with greater CAC burden (β 0.38, 95% credible interval 0.15 - 0.61).

**Conclusions:** Polygenic risk for angiographic coronary disease burden is reflected in clinically reported coronary CTA severity measures, including CAD-RADS and CAC. These findings demonstrate that inherited susceptibility to CAD manifests as greater anatomic atherosclerotic burden at the time of clinical presentation and support further investigation of genetic risk integration into imaging-based cardiovascular risk assessment.

---

Conventional cardiovascular risk assessment relies heavily on age-weighted clinical risk factors and underestimates risk in younger individuals, in whom substantial coronary atherosclerotic burden may be present despite low estimated short-term risk ^1^. Coronary computed tomography angiography (CTA) provides direct, noninvasive assessment of coronary atherosclerosis and is increasingly used in the evaluation of patients with stable chest pain and other symptoms suggestive of CAD ^2^.

Polygenic risk scores (PRS) for CAD capture inherited susceptibility to atherosclerosis across the life course and have been shown to be associated with CAD events and measures of anatomic disease ^3^. Prior studies have demonstrated associations between genetic risk and coronary plaque burden, progression, and high-risk plaque features in selected CTA cohorts, as well as with advanced atherosclerotic pathology in autopsy series ^4^. More recently, PRS trained specifically on angiographic coronary disease burden have been described, offering a genetic construct more closely aligned with anatomic severity rather than binary disease status ^5^. However, the extent to which inherited genetic predisposition translates into clinically reported coronary CTA severity categories, including Coronary Artery Disease-Reporting and Data System (CAD-RADS) and coronary artery calcium (CAC) scores, is unknown.

In this study, we leveraged the Penn Medicine BioBank (https://pmbb.med.upenn.edu/), an electronic health record-linked biobank with substantial ancestral diversity, to examine the relationship between polygenic risk for angiographic coronary disease burden^5^ and coronary CTA findings in a real-world clinical setting. This study was approved by the institutional review board of the University of Pennsylvania with waiver of informed consent. Using *pgsc_calc* (https://pgsc-calc.readthedocs.io/en/latest/), we calculated a previously published genome-wide PRS for angiographic CAD burden among PMBB participants as previously described^5^. Using the open-source large language model Llama 3.1 8B (Meta Platforms, Inc.) with temperature set to 0, we extracted CAD-RADS scores and CAC information from clinically obtained coronary CTA radiology reports, enabling scalable phenotyping directly from existing clinical reports. Compared to clinician-extracted values from 30 randomly selected reports blindly reviewed by a radiologist (KH), the LLM achieved multiclass precision for CAD-RADS classification of 0.90, recall of 0.94 and kappa 0.98; extracted CAC values demonstrated near-perfect agreement with manual abstraction with Spearman ρ = 0.999 (p < 0.001 for all).

Among 630 PMBB participants with available genotypes and CAD-RADS (n = 599) or CAC (n = 367) from coronary CTA reports, the median age was 59 years (IQR = 49-68), 53% were female, 62% were genetically similar to a European reference population and 34% were genetically similar to an African reference population. Higher polygenic risk for angiographic coronary disease burden was associated with more severe coronary atherosclerosis on CTA, as reflected by CAD-RADS category (Figure 1). In an ordinal cumulative logit model, each 1-SD higher PRS was associated with a 20% higher odds of belonging to a more severe CAD-RADS category (cumulative OR 1.2, 95% credible interval 1.06–1.44), independent of age and sex. This relationship was observed across age and sex strata despite lower absolute disease burden among younger individuals and women, indicating that genetic risk influences disease severity independent of baseline demographic risk. Similarly, higher polygenic risk was significantly associated with greater CAC. Each 1-SD higher PRS corresponded to a 48% higher CAC burden (beta 0.38, 95% credible interval 0.15–0.61), equivalent to approximately 5–6 years of aging in terms of CAC burden. These findings demonstrate that polygenic risk is reflected in the anatomic burden of coronary atherosclerosis, including both stenotic severity and calcified plaque, at the time of clinical presentation.

**Figure 1.**
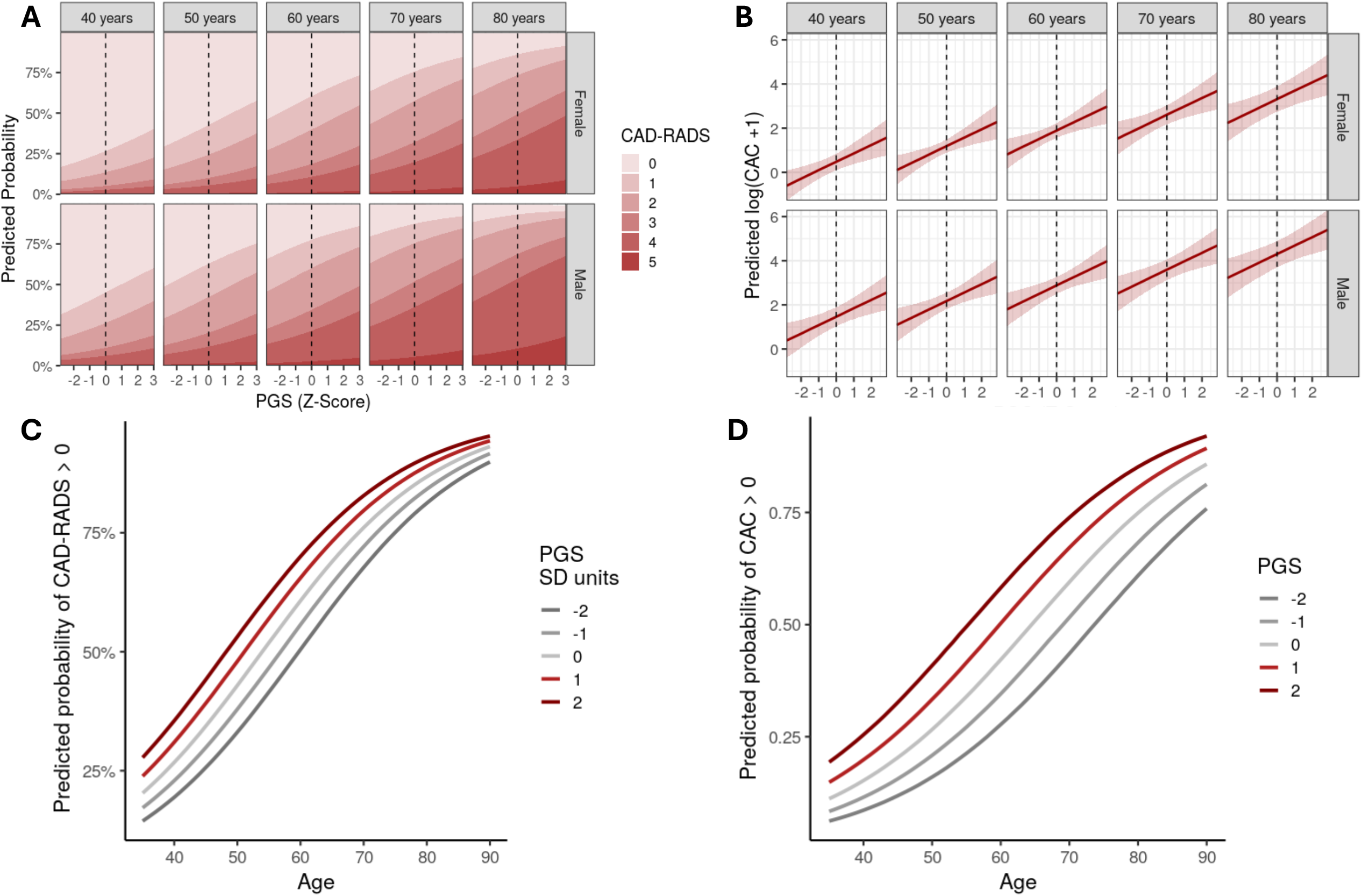
Association of polygenic risk for angiographic coronary disease burden with coronary CTA severity in routine clinical practice. We evaluated associations between polygenic risk and CTA-derived measures of coronary disease severity across age and sex strata among individuals undergoing coronary CTA for clinical indications. (A) Predicted probabilities of CAD-RADS categories (0–5) and (B) predicted CAC across the distribution of standardized polygenic risk score (PGS Z-score), stratified by age and sex. (C) Predicted probability of CAD-RADS >0 and (D) coronary artery calcium (CAC) >0 by age for representative PGS values ranging from −2 to +2 standard deviations from the mean. Predictions were obtained from Bayesian cumulative ordinal logit regression (CAD-RADS) and Bayesian linear regression of log-transformed CAC (log[CAC+1]) implemented in *brms*, using Hamiltonian Monte Carlo sampling. Posterior expected probabilities (CAD-RADS) and posterior expected transformed values (CAC) were computed across PRS values while conditioning on fixed age and sex. Shaded regions in B represent 95% posterior credible intervals.

Our study population represents individuals referred for coronary CTA in routine clinical care, frequently to exclude obstructive coronary artery disease. As such, this is not a screening cohort, and referral patterns, symptom burden, and clinician decision-making likely influence both imaging and disease prevalence. These factors introduce selection and collider biases that may attenuate or distort associations relative to unselected populations. Nevertheless, the persistence of graded associations with both CAD-RADS and CAC within this clinically enriched setting underscores the robustness of inherited risk as a contributor to anatomic disease severity. By linking inherited risk to routinely reported imaging phenotypes at the point of clinical evaluation, this work provides a foundation for understanding how genetic susceptibility manifests in practice and informs future studies assessing whether genetic risk might help guide imaging-based strategies in broader, preclinical populations.

## Data Availability

Raw data for the analysis dataset are not publicly available to preserve individuals privacy per the Health Insurance Portability and Accountability Act Privacy Rule

## Acknowledgements

Penn Medicine Biobank Banner Author List and Contribution Statements PMBB Leadership Team

Daniel J. Rader, M.D.; Marylyn D. Ritchie, Ph.D.

Contribution: All authors contributed to securing funding, study design, and oversight. All authors reviewed the final version of the manuscript.

## Patient Recruitment and Regulatory Oversight

JoEllen Weaver; Nawar Naseer, Ph.D., M.P.H.; Giorgio Sirugo, M.D., Ph.D.; Afiya Poindexter; Jenna Dever; Aidan Harvey; Sydney Linn; Naman Srivastava

Contributions: JW manages participant recruitment and regulatory oversight. NN manages participant engagement, assists with regulatory oversight, and supports researcher access. GS supports researcher access. AP, JD, AH, SL, and NS perform recruitment and enrollment of participants.

## Lab Operations

JoEllen Weaver; Meghan Livingstone; Fred Vadivieso; Stephanie DerOhannessian; Teo Tran; Julia Stephanowski; Salma Santos; Ned Haubein, Ph.D.; Joseph Dunn

Contribution: JW, ML, FV, and SD oversee lab operations. ML, FV, AK, SD, TT, JS, and SS perform sample processing. NH and JD manage sample tracking and the laboratory information management system.

## Clinical Informatics

Anurag Verma, Ph.D.; Colleen Morse Kripke, M.S., DPT, MSA; Marjorie Risman, M.S.; Renae Judy, B.S.; Colin Wollack, M.S.

Contribution: All authors contributed to the development and validation of clinical phenotypes used to identify study participants and (when applicable) controls.

## Genome Informatics

Anurag Verma, Ph.D.; Shefali S. Verma, Ph.D.; Scott Damrauer, M.D.; Yuki Bradford, M.S.; Scott Dudek, M.S.; Theodore Drivas, M.D., Ph.D.; Zachary Rodriguez, Ph.D.

Contribution: AV, SSV, and SD developed the analysis, design, and infrastructure needed for quality control of genotype and exome data. YB performed analyses. TD and AV provided variant and gene annotations and interpreted variant function.

## References

1. An, J. et al. Incidence of Atherosclerotic Cardiovascular Disease in Young Adults at Low Short-Term But High Long-Term Risk. J Am Coll Cardiol 81, 623–632 (2023).

2. Danzi, G. B. & Piccolo R. CT or Invasive Coronary Angiography in Stable Chest Pain. N Engl J Med 387, 378 (2022).

3. Kullo, I. J. Clinical use of polygenic risk scores: current status, barriers and future directions. Nat Rev Genet 27, 246–263 (2026).

4. Nurmohamed, N. S. et al. Polygenic Risk Is Associated With Long-Term Coronary Plaque Progression and High-Risk Plaque. JACC Cardiovasc Imaging 17, 1445–1459 (2024).

5. Tsao, N. L. et al. Angiographic Burden of Coronary Atherosclerosis Partially Mediates the Association Between ASCVD Risk Factors and Outcomes. Circ Genom Precis Med e005266 (2026) doi:10.1161/CIRCGEN.125.005266.

